# Investigating Genetic and Familial Risks in Childhood ALL: A Longitudinal Virtual Study Using aiHumanoid Simulations of TEL-AML1 Gene Fusion

**DOI:** 10.1101/2024.06.19.24309162

**Authors:** WR Danter

**Affiliations:** Windsor, Canada

**Keywords:** Childhood cancer, Acute Lymphoblastic Leukemia (ALL), TEL-AML1 (ETV6-RUNX1) gene fusion, family history (FHx), aiHumanoid, DeepNEU, AI simulations

## Abstract

Acute Lymphoblastic Leukemia (ALL) is the most common childhood cancer, presenting significant challenges in early diagnosis and effective treatment. Recent advances in gene profiling have identified a pivotal role for the TEL-AML1 (ETV6-RUNX1) gene fusion, present in about one quarter of pediatric ALL cases. This gene fusion is often associated with a more benign course and requires additional genetic abnormalities known as *Second Hits*, to cause overt leukemia. Our current study utilizes Fuzzy Cognitive Maps (FCMs) to model the intricate genetic interplays and potential progression paths of ALL in children, focusing on those with the TEL-AML1 gene fusion within a familial leukemia context.

In this virtual longitudinal study, we leverage cohorts of aiHumanoid simulations to explore the foundational role of the TEL-AML1 fusion gene in the genesis of ALL, examining its impact when combined with a family history of leukemia. The simulations predict how genetic and environmental factors might influence disease onset and progression, providing a platform for early diagnosis and progression monitoring.

Our findings suggest that family history significantly increases the risk and modifies the disease course in carriers of the TEL-AML1 fusion gene, indicating a need for targeted surveillance and potential early interventions in these high-risk groups.

## Introduction

Acute Lymphoblastic Leukemia (ALL) is the most common form of leukemia found in children, accounting for about 30 percent of all pediatric cancers. It is also the most prevalent cancer among children and adolescents in the United States, representing 20% of all cancers diagnosed in people less than 20 years of age. ALL represents approximately (i) 25% of cancer diagnoses and (ii) 75% of all leukemias, in children younger than 15 years [1,2,3]. This blood cancer is characterized by the uncontrolled proliferation of immature white blood cells in the bone marrow, leading to significant hematological disruptions and systemic complications.

Genetic research over the last decade has significantly unraveled the complexities of the disease, highlighting several key genetic and molecular drivers behind ALL. Notably, the TEL-AML1 (ETV6-RUNX1) fusion gene has emerged as a critical factor, which can be detected in approximately one quarter of all pediatric ALL cases. This gene fusion, resulting from a chromosomal translocation, is often associated with a favorable prognosis but requires additional genetic mutations — known as *Second Hits* — to cause overt leukemia [4,5].

The “Second Hit” theory suggests that these subsequent genetic events are necessary for the clonal expansion of leukemic cells, providing a framework for understanding the multi-step nature of leukemia oncogenesis [6]. For instance, mutations in tumor suppressor genes or additional oncogenic events like loss of homogeneity (LOH), can fulfill the role of a second hit, exacerbating the leukemic process initiated by a first hit like the TEL-AML1 gene fusion.

AI systems based on Fuzzy Cognitive Maps (FCMs) offer an innovative approach to model these complex interactions within biological systems. FCMs utilize fuzzy or grey scale logic to map causal relationships and their strength between various components of a system, such as genetic factors, environmental influences, and cellular pathways. This modeling technique which most resembles fully connected, recurrent artificial neural networks (RNN) has been successfully applied in various fields of medicine and is particularly suited for capturing the dynamic and multifactorial nature of diseases like ALL [7,8,9,10].

In this virtual study, we develop a comprehensive FCM simulations for modelling the intricate network of genetic interactions in childhood ALL, with a particular focus on the impact of the TEL-AML1 fusion gene alone and in the setting of a family history of leukemia regarding early diagnosis, disease development and progression.

This AI guided approach may be particularly relevant for specific difficult to treat gene mutation profiles in childhood ALL.

## Methods

### Overview

The current study utilizes aiHumanoid simulations as virtual subjects in a longitudinal investigation into the development of acute lymphoblastic leukemia (ALL) in childhood. We aim to better understand the importance of a family history of leukemia with and without a TEL-AML1 gene fusion and the risk of developing ALL. Part 1 of this project will focus on the preleukemic state, early diagnosis, and disease progression.

1. Updating the aiHumanoid Simulation to v8.4:

The previous version 8.3 of the aiHumanoid [9,10] underwent revisions to v8.4. The main differences are that the revised version integrates new simulations for specific ALL associated mutations and an updated subsystem for the diagnosis of ALL in children. The number of integrated organoid simulations remains at 21. A literature validation of the ALL simulations, employing the same approach used in previous versions was applied to the updated simulations comprising v8.4.

2. ALL Validation Profile in the aiHumanoid Simulations:

To confirm a diagnosis of ALL in the affected aiHumanoid simulated children, a list of twenty genotypic and phenotypic features was assembled from the literature for evaluation and are presented in Appendix A. All features were statistically significantly different from controls for multiple age matched cohorts and regarding the diagnosis of ALL. The present analysis employed a combination of the nonparametric Wilcoxon signed rank test and the Cliff’s delta effect size estimates.

3. Study Design and Objectives:

This project is our most recent virtual longitudinal study using the aiHumanoid simulations. The objectives of this study were: (i) to evaluate the impact of a family history of any leukemia with or without the presence of a TEL-AML1 gene fusion on the risk of developing ALL in childhood.(ii) to evaluate a panel of disease features for the purpose of identifying opportunities for early diagnosis, and (iii) to better understand ALL development and progression from birth to age 20 years.

### The Virtual subjects used in this study

The profiles for twenty-five unique and healthy virtual subjects were synthesized by GPT-4, an advanced large language model (December 2023 version,) at https://chat.openai.com/). GPT4 used its extensive database encompassing medical literature, patient profiles, and related clinical information, to synthesize diverse and representative draft profiles for twenty-five healthy children. Each subject profile was reviewed by an experienced physician prior to enrolment. This longitudinal study design permitted us to create a virtual study with 4 genotype groups (WT, WT plus a family history of leukemia at any age, TEL-AML1 gene fusion and TEL-AML1 fusion in the presence of a family history of leukemia at any age) X 6 (age groups) X 25 (virtual subjects), the equivalent of data from 600 young patients. The virtual subjects used in this study serve as hypothetical, but commonly encountered population based examples of risks associated with the development of ALL in children in specific affected cohorts but do not represent actual individuals or precise medical histories.

Inclusion Criteria (WT/Healthy cohorts):

1. Generally healthy at birth
2. Ages Birth (0 years) to 20 years
3. Approximately equal representation of boys and girls
4. All required individual data are available

Exclusion criteria:

5. Age greater than 20 years of age
6. Any documented preexisting genetic abnormalities
7. Any documented disease processes prior to or at birth

### The Affected Subjects

The ALL associated risk factors studied included: (i) the WT state plus a FHx of leukemia, (ii) the TEL-AML1 gene fusion and (iii) the TEL-AML1 gene fusion in the context of a family history of leukemia. The six age cohorts of twenty-five healthy subjects each underwent AI gene editing to introduce loss of function (LOF) mutations for each of the two ALL associated TEL-AML1 fusion studied (11). This process created twenty-four highly matched cohorts where the only difference was the presence or absence of a specific genotype. In these well-matched cohorts, properties like obesity, hypertension and Type 2 Diabetes are emergent properties primarily associated with age. The virtual approach has the major advantage that all subjects’ data were available for analysis since there was no attrition which would be common in traditional longitudinal study of this kind. The data from all cohorts were evaluated beginning at birth (0 years) and continuing at 5-year intervals up to and including age 20 years of age (6 age cohorts).

### Statistical Analysis

The Null hypothesis states that there are no statistically significant differences or at least medium effect sizes for the three affected groups compared to the age matched wild type (WT) subjects. The data was not normally distributed, so the non-parametric Wilcoxon signed rank test was used. Given that multiple tests (N=20) were conducted, the conservative Bonferroni correction was applied. The corrected p value to achieve significance therefore became 0.05/20 < 0.0025 for this study.

The alternative hypothesis states that there are significant differences in the three affected groups compared to the healthy controls (WT).

The true effect size was estimated using Cliff’s delta. Cliff’s delta was used because the data were not normally distributed with a sample size of twenty-five subjects per cohort. To calculate Cliff’s delta the continuous data was transformed into interval data based on whether the data from the affected group was larger or smaller than the unaffected group. The cliff’s delta was then calculated as (N (larger than) – N (smaller than))/the standard deviation of the differences between the groups (12,13). This produced a range of effect size estimates between −1 (a negative effect) and +1 (a positive effect). A value close to zero was interpreted as having no effect. To determine the size of the effect we used the following heuristic scale: d <0.147 (negligible), d = 0.147 to <0.330 (small), d = 0.330 to <0.474 (medium) and d >= 0.474 (large) as suggested in (14). To compensate for the modest sample size per cohort, all Cliff’s d values were modified using the Hedges correction (15) which was calculated to be 0.984. Final effect size estimates were obtained by multiplying the initial effect sizes by 0.984. The 95% CI around the effect size estimate was calculated using the SE of the differences/square root of the sample size (2N data points).

## Results

In the present study, v8.4 (2024) of the DeepNEU database, which is characterized by modest upgrades to its previous version 8.3, was used [18]. Specifically, v8.3 had 7447 concepts and 69581 nonzero causal relationships. Version 8.4 has 7529 concepts and 70587 relationships. This means that for every nonzero concept in the causal relationship matrix, there are approximately 9.4 incoming and outgoing causal relationships.

Furthermore, v8.4 contains new validated simulations for the diagnosis of acute lymphoblastic leukemia (ALL) in children and leukemia associated pathways. The total number of integrated organoid simulations is unchanged at 21. The previously implemented early stopping algorithm (v8.2) was retained in this updated version. For the purposes of this longitudinal study, we chose to use the results after 14 iterations, as determined through a three valued moving average, which minimized any evidence of overfitting.

1. Family History (FHx) of Leukemia (Any): These data are summarized in Table 1

**Table 1:** Summary of the Cliff’s delta (d) effect size data for the combination of Wild Type Plus Family Hixtory of Leukemia across all six Age cohorts.

| Age | TEM-AML1 Fusio | Splenomegaly | Anemia | Fever | CD10/Neprilysin | CD19 | CD2 | CD20 | CD22 | CD3 | CD4 | CD45 |
| --- | --- | --- | --- | --- | --- | --- | --- | --- | --- | --- | --- | --- |
| 0 | 0.000 | 0.984 | 0.000 | 0.000 | -0.236 | 0.276 | 0.945 | -0.197 | -0.905 | -0.984 | 0.118 | 0.000 |
| 2-3 | 0.000 | 0.984 | -0.039 | 0.000 | -0.079 | 0.157 | 0.905 | -0.905 | -0.984 | -0.984 | 0.276 | 0.000 |
| 5 | 0.000 | 0.984 | -0.039 | 0.000 | -0.433 | 0.197 | 0.354 | -0.433 | -0.984 | -0.984 | -0.276 | 0.000 |
| 10 | 0.000 | 0.984 | -0.039 | 0.000 | -0.551 | 0.394 | 0.157 | -0.118 | -0.905 | -0.984 | 0.354 | 0.000 |
| 15 | 0.000 | 0.984 | -0.039 | 0.000 | -0.590 | 0.512 | 0.039 | 0.512 | -0.039 | -0.905 | -0.118 | -0.394 |
| 20 | 0.000 | 0.984 | -0.039 | 0.000 | -0.630 | 0.984 | 0.039 | 0.197 | 0.905 | -0.669 | 0.039 | -0.039 |
| Average | 0.000 | 0.984 | -0.033 | 0.000 | -0.420 | 0.420 | 0.407 | -0.157 | -0.485 | -0.918 | 0.066 | -0.072 |
| +/- 95% CI | 0.000 | 0.000 | 0.013 | 0.000 | 0.176 | 0.244 | 0.334 | 0.394 | 0.618 | 0.101 | 0.190 | 0.127 |
| Scale | -1 | -0.5 | 0 |  | 0.50 | 1 |  |  |  |  |  |  |
| Age | B Cell<br>CD79a | T Cell<br>CD8 | IgA | IgG | B Cell<br>IgM | LDH | LeukocytesWBC | Lymphoblasts | MPO | Platelets | T Cell<br>T Cell Receptor | B Cell<br>TdT |
| 0 | -0.984 | -0.748 | -0.984 | -0.984 | -0.984 | 0.984 | -0.984 | 0.984 | 0.000 | -0.984 | -0.827 | 0.984 |
| 2-3 | -0.905 | -0.079 | -0.354 | -0.905 | -0.905 | 0.984 | -0.984 | 0.905 | 0.000 | -0.827 | -0.276 | 0.905 |
| 5 | -0.905 | 0.748 | -0.197 | -0.905 | -0.905 | 0.118 | -0.984 | 0.905 | 0.000 | -0.905 | -0.354 | 0.905 |
| 10 | -0.905 | -0.236 | -0.905 | -0.905 | -0.905 | -0.276 | -0.984 | 0.905 | 0.000 | -0.905 | 0.039 | 0.905 |
| 15 | -0.905 | -0.039 | -0.905 | -0.905 | -0.905 | 0.118 | -0.984 | 0.905 | 0.000 | -0.905 | -0.118 | 0.905 |
| 20 | -0.827 | -0.433 | 0.118 | -0.905 | -0.905 | 0.118 | -0.905 | 0.905 | 0.000 | -0.905 | -0.905 | 0.905 |
| Average | -0.905 | -0.131 | -0.538 | -0.918 | -0.918 | 0.341 | -0.971 | 0.918 | 0.000 | -0.905 | -0.407 | 0.918 |
| +/- 95% CI | 0.040 | 0.403 | 0.367 | 0.026 | 0.026 | 0.417 | 0.026 | 0.026 | 0.000 | 0.040 | 0.305 | 0.026 |

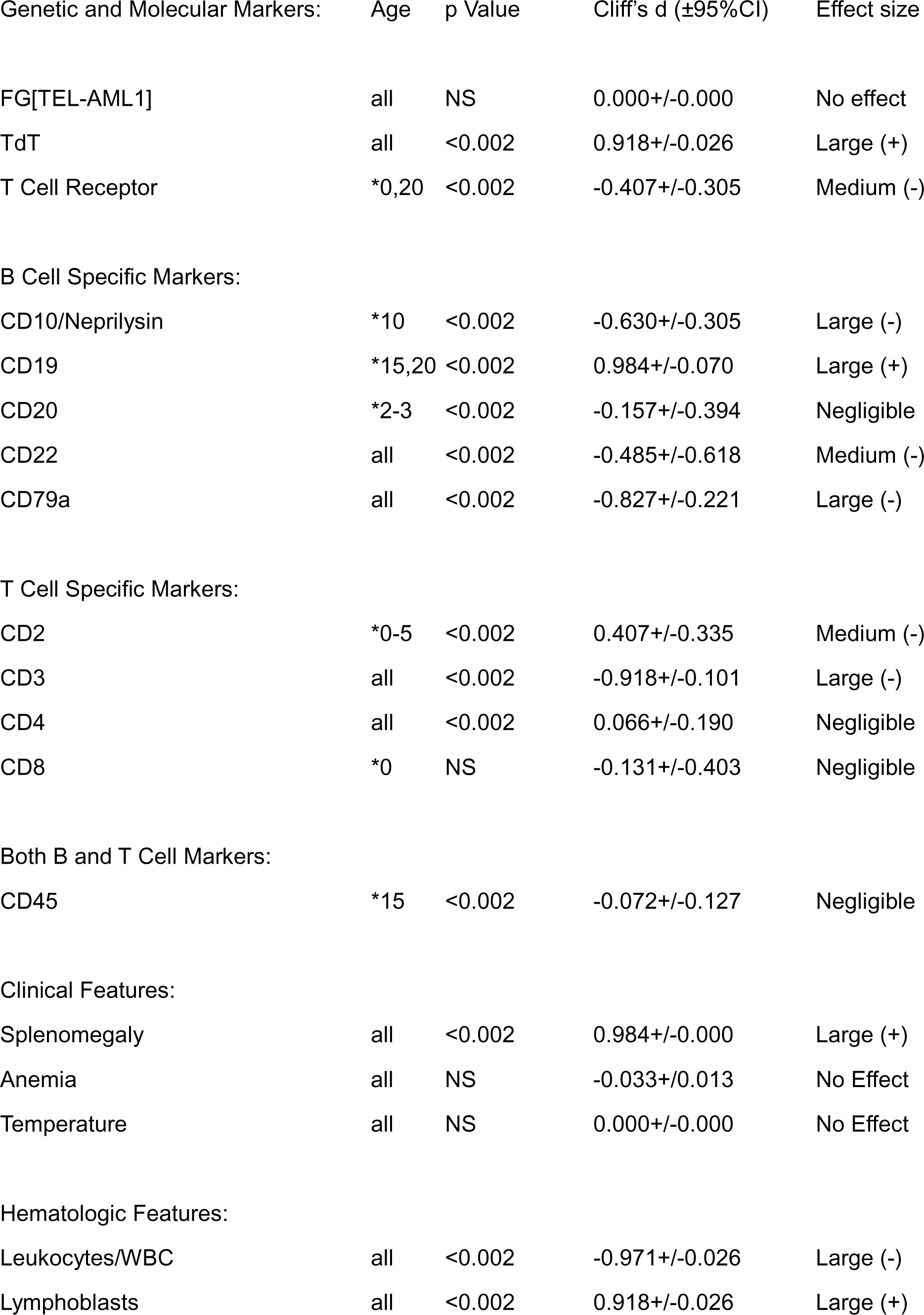
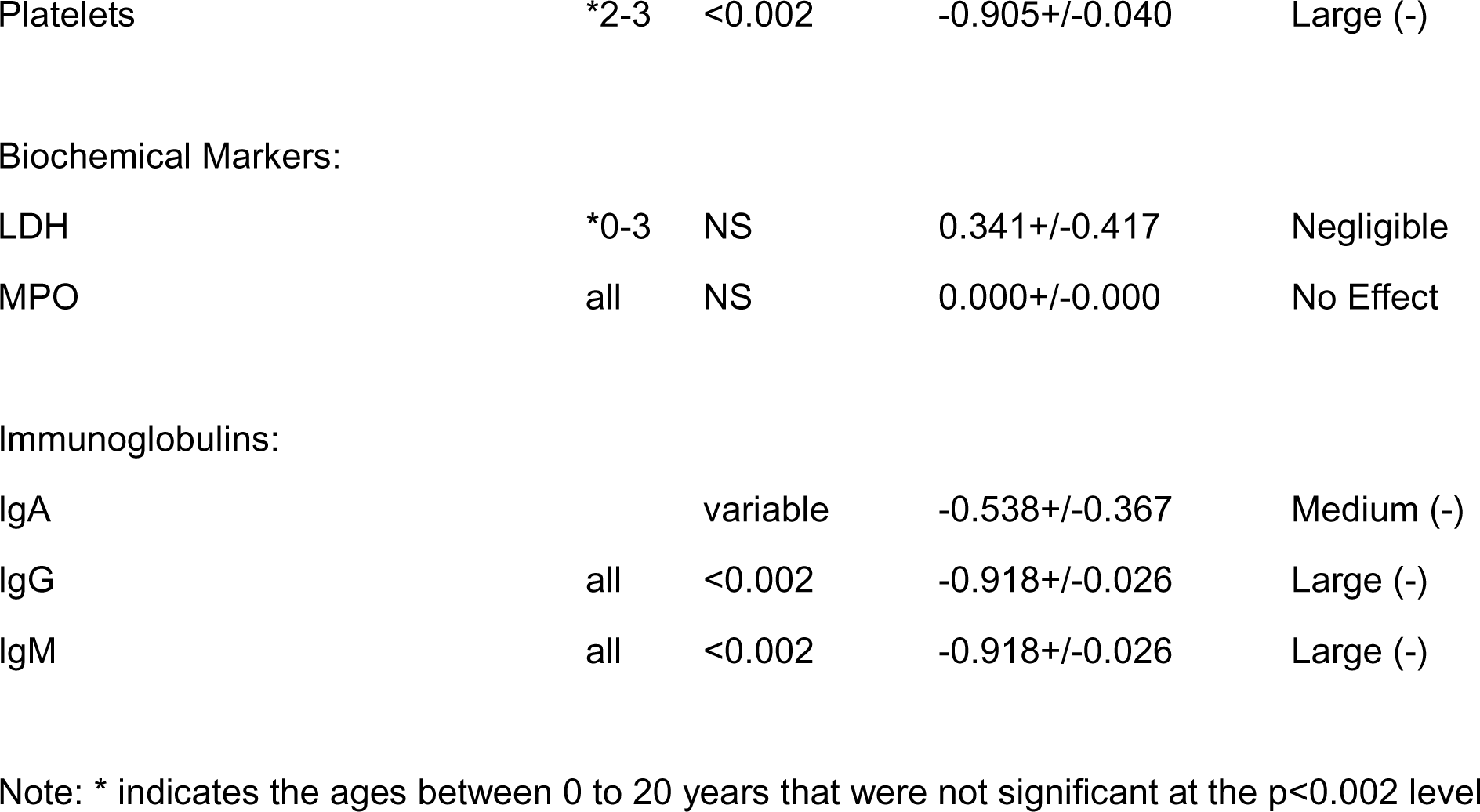

| Genetic and Molecular Markers: | Age | p Value | Cliff's d ($\pm 95\%CI$) | Effect size |
| --- | --- | --- | --- | --- |
| FG[TEL-AML1] | all | NS | 0.000+/-0.000 | No effect |
| TdT | all | <0.002 | 0.918+/-0.026 | Large (+) |
| T Cell Receptor | *0,20 | <0.002 | -0.407+/-0.305 | Medium (-) |
| B Cell Specific Markers: |  |  |  |  |
| CD10/Neprilysin | *10 | <0.002 | -0.630+/-0.305 | Large (-) |
| CD19 | *15,20 | <0.002 | 0.984+/-0.070 | Large (+) |
| CD20 | *2-3 | <0.002 | -0.157+/-0.394 | Negligible |
| CD22 | all | <0.002 | -0.485+/-0.618 | Medium (-) |
| CD79a | all | <0.002 | -0.827+/-0.221 | Large (-) |
| T Cell Specific Markers: |  |  |  |  |
| CD2 | *0-5 | <0.002 | 0.407+/-0.335 | Medium (-) |
| CD3 | all | <0.002 | -0.918+/-0.101 | Large (-) |
| CD4 | all | <0.002 | 0.066+/-0.190 | Negligible |
| CD8 | *0 | NS | -0.131+/-0.403 | Negligible |
| Both B and T Cell Markers: |  |  |  |  |
| CD45 | *15 | <0.002 | -0.072+/-0.127 | Negligible |
| Clinical Features: |  |  |  |  |
| Splenomegaly | all | <0.002 | 0.984+/-0.000 | Large (+) |
| Anemia | all | NS | -0.033+/-0.013 | No Effect |
| Temperature | all | NS | 0.000+/-0.000 | No Effect |
| Hematologic Features: |  |  |  |  |
| Leukocytes/WBC | all | <0.002 | -0.971+/-0.026 | Large (-) |
| Lymphoblasts | all | <0.002 | 0.918+/-0.026 | Large (+) |
| Platelets | *2-3 | <0.002 | -0.905+/-0.040 | Large (-) |
| Biochemical Markers: |  |  |  |  |
| LDH | *0-3 | NS | 0.341+/-0.417 | Negligible |
| MPO | all | NS | 0.000+/-0.000 | No Effect |
| Immunoglobulins: |  |  |  |  |
| IgA |  | variable | -0.538+/-0.367 | Medium (-) |
| IgG | all | <0.002 | -0.918+/-0.026 | Large (-) |
| IgM | all | <0.002 | -0.918+/-0.026 | Large (-) |

Note: \* indicates the ages between 0 to 20 years that were not significant at the $p < 0.002$ level

### Summary

Children with a family history (FHx) of leukemia and no other known risk factors exhibit significant hematologic and immunologic changes when compared to the wild-type (WT) state. Notable findings include a marked increase in splenomegaly (Cliff’s delta = 0.984, p < 0.002) and lymphoblasts (Cliff’s delta = 0.918, p < 0.002), indicating large effect sizes. Additionally, B-cell activity is heightened, as evidenced by the significant increase in CD19 (Cliff’s delta = 0.984, p < 0.002). Conversely, there are significant decreases in leukocyte counts (Cliff’s delta = −0.971, p < 0.002), platelets (Cliff’s delta = −0.905, p < 0.002), and immunoglobulins IgG and IgM (both with Cliff’s delta = −0.918, p < 0.002), indicating large negative effects.

Other notable changes include a moderate decrease in CD10/Neprilysin (Cliff’s delta = −0.630, p < 0.002) and CD79a (Cliff’s delta = −0.827, p < 0.002), as well as a large decrease in the T-cell marker CD3 (Cliff’s delta = −0.918, p < 0.002). The overall pattern of increased immature lymphoid cells and decreased functional leukocytes and platelets suggests a predisposition to hematologic abnormalities. These findings underscore the significant impact of familial predisposition on hematologic and immunologic parameters in children, highlighting the need for close monitoring and early intervention in those with a family history of leukemia.

The Link between a Family History of Leukemia and Early Childhood Hematologic Changes

The observed data align with known early indicators and risk factors for developing B-cell Acute Lymphoblastic Leukemia (ALL). While these children do not have B-cell ALL, the significant increase in lymphoblasts (Cliff’s delta = 0.918) and splenomegaly (Cliff’s delta = 0.984) are consistent with early leukemic changes (16,17,18,19). The heightened B-cell activity, as evidenced by increased CD19 levels, mirrors the overproduction of B-cells characteristic of B-cell ALL (18,19). Decreases in leukocyte counts, platelets, and immunoglobulins suggest early bone marrow stress and immune changes, which can precede the onset of leukemia (18,19). These data indicate a potential early disruption in hematologic and immunologic parameters, emphasizing the importance of vigilant monitoring and preventive care to mitigate the risk of progression to B-cell ALL in children with a family history of any type of leukemia (16,17,18,19).

2. TEL-AML1 Fusion: These data are summarized in Table 2

**Table 2:** Summary of the Cliff’s delta (d) effect size data for the presence of a TEM-AML1 gene fusion mutation across all six Age cohorts.

| Age | TEM-AML1 Fusion | Splenomegaly | Anemia | Fever | CD10/Neprilysin | CD19 | CD2 | CD20 | CD22 | CD3 | CD4 | CD45 |
| --- | --- | --- | --- | --- | --- | --- | --- | --- | --- | --- | --- | --- |
| 0.000 | 0.984 | 0.984 | 0.984 | 0.000 | 0.984 | 0.984 | 0.039 | 0.984 | 0.984 | -0.984 | -0.905 | 0.039 |
| 2-3 | 0.984 | 0.984 | 0.945 | 0.000 | 0.984 | 0.984 | 0.039 | 0.984 | 0.984 | -0.984 | -0.984 | 0.039 |
| 5.000 | 0.984 | 0.984 | 0.945 | 0.000 | 0.984 | 0.984 | 0.039 | 0.984 | 0.984 | -0.984 | -0.984 | -0.354 |
| 10.000 | 0.984 | 0.984 | 0.945 | 0.000 | 0.984 | 0.984 | 0.039 | 0.827 | 0.984 | -0.039 | -0.984 | 0.000 |
| 15.000 | 0.984 | 0.984 | 0.945 | 0.000 | 0.984 | 0.984 | 0.039 | 0.984 | 0.984 | -0.039 | -0.276 | -0.394 |
| 20.000 | 0.984 | 0.984 | 0.945 | 0.000 | 0.984 | 0.984 | 0.039 | 0.984 | 0.984 | -0.039 | -0.905 | -0.551 |
| Average | 0.984 | 0.984 | 0.951 | 0.000 | 0.984 | 0.984 | 0.039 | 0.958 | 0.984 | -0.512 | -0.840 | -0.203 |
| 95% CI | 0.000 | 0.000 | 0.013 | 0.000 | 0.000 | 0.000 | 0.000 | 0.051 | 0.000 | 0.414 | 0.223 | 0.208 |
| Scale | -1.000 | -0.500 | 0.000 |  | 0.500 | 1.000 |  |  |  |  |  |  |
| Age | CD79a | CD8 | IgA | IgG | IgM | LDH | LeukocytesWBC | Lymphoblasts | MPO | Platelets | T Cell Receptor | TdT |
| 0.000 | 0.905 | -0.039 | 0.905 | -0.984 | -0.984 | 0.984 | -0.984 | 0.984 | 0.512 | -0.118 | 0.039 | 0.984 |
| 2-3 | 0.905 | 0.748 | 0.984 | -0.984 | -0.905 | 0.984 | -0.984 | 0.984 | 0.984 | -0.905 | 0.039 | 0.984 |
| 5.000 | 0.905 | 0.433 | 0.905 | -0.984 | -0.905 | 0.984 | -0.984 | 0.984 | 0.551 | -0.039 | 0.039 | 0.984 |
| 10.000 | -0.748 | -0.039 | 0.905 | -0.984 | -0.905 | 0.984 | -0.984 | 0.984 | 0.512 | -0.197 | -0.905 | 0.984 |
| 15.000 | -0.905 | 0.039 | 0.984 | -0.984 | -0.905 | 0.984 | -0.984 | 0.984 | 0.551 | -0.827 | 0.039 | 0.984 |
| 20.000 | -0.748 | -0.039 | 0.984 | -0.984 | -0.905 | 0.984 | -0.984 | 0.984 | 0.512 | 0.590 | 0.039 | 0.984 |
| Average | 0.052 | 0.184 | 0.945 | -0.984 | -0.918 | 0.984 | -0.984 | 0.984 | 0.604 | -0.249 | -0.118 | 0.984 |
| 95% CI | 0.749 | 0.265 | 0.034 | 0.000 | 0.026 | 0.000 | 0.000 | 0.000 | 0.150 | 0.443 | 0.309 | 0.000 |

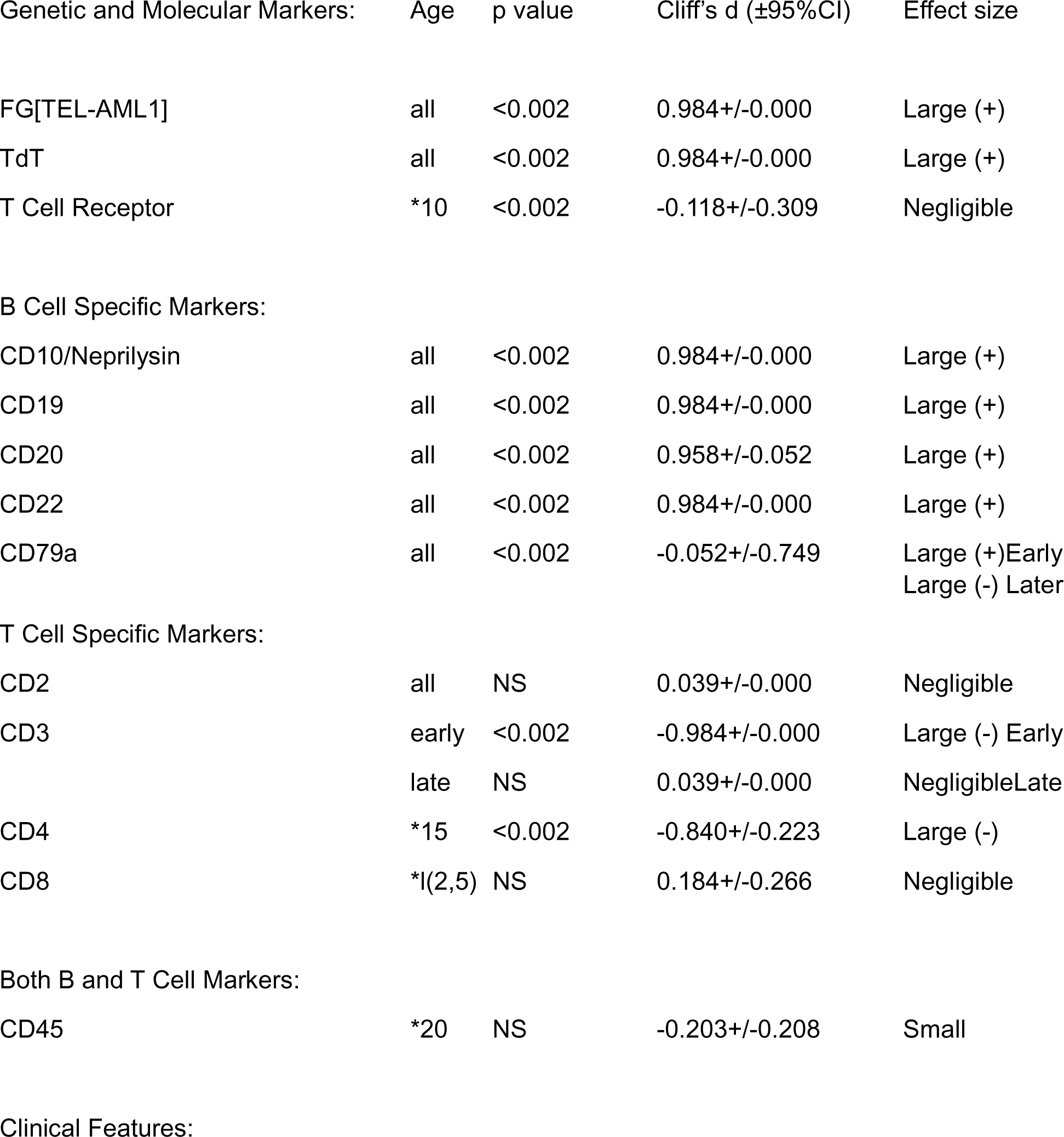
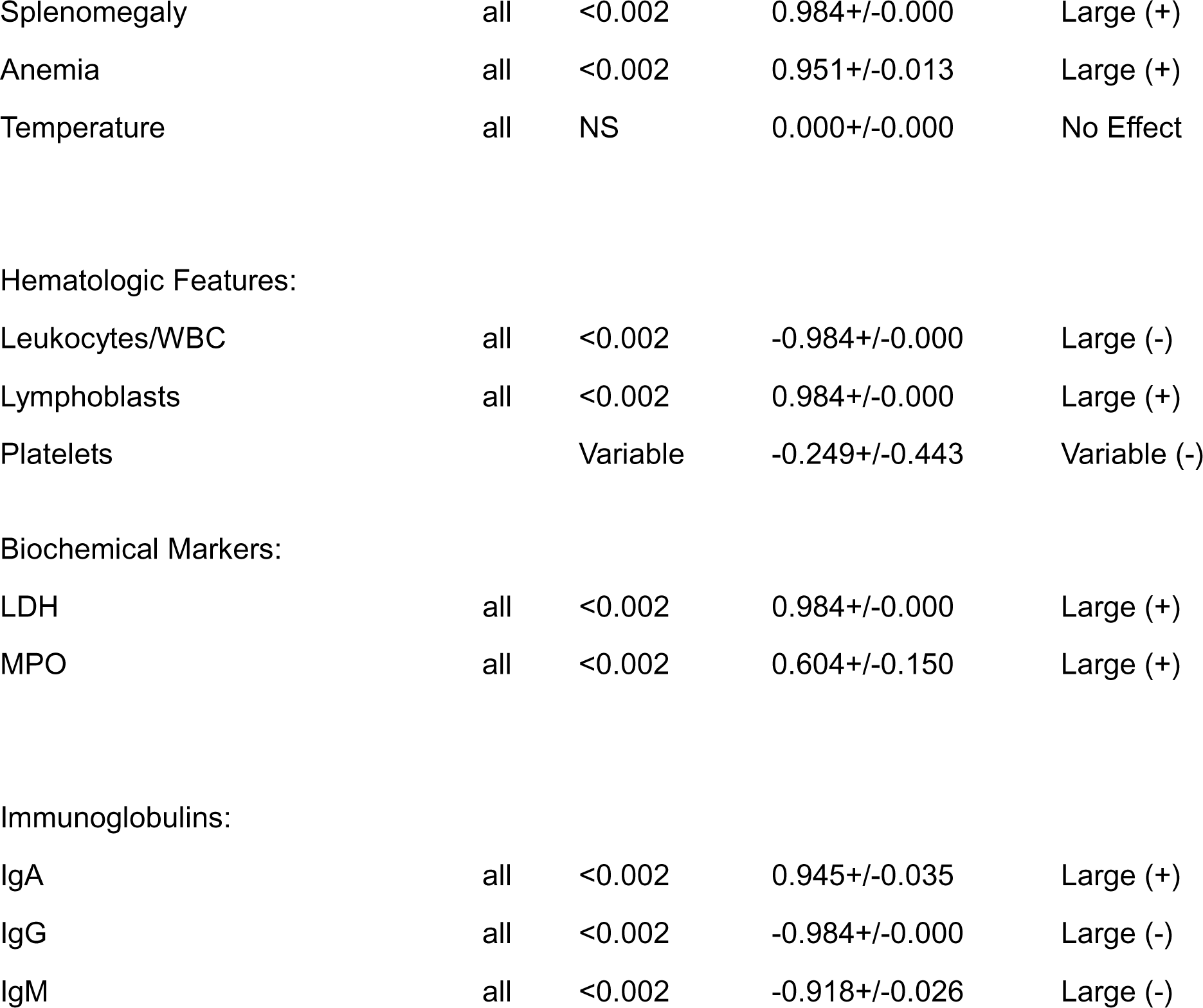

2. TEL-AML1 Fusion: These data are summarized in Table 2
| Genetic and Molecular Markers: | Age | p value | Cliff's d ($\pm 95\%CI$) | Effect size |
| --- | --- | --- | --- | --- |
| FG[TEL-AML1] | all | <0.002 | 0.984+/-0.000 | Large (+) |
| TdT | all | <0.002 | 0.984+/-0.000 | Large (+) |
| T Cell Receptor | *10 | <0.002 | -0.118+/-0.309 | Negligible |
| B Cell Specific Markers: |  |  |  |  |
| CD10/Neprilysin | all | <0.002 | 0.984+/-0.000 | Large (+) |
| CD19 | all | <0.002 | 0.984+/-0.000 | Large (+) |
| CD20 | all | <0.002 | 0.958+/-0.052 | Large (+) |
| CD22 | all | <0.002 | 0.984+/-0.000 | Large (+) |
| CD79a | all | <0.002 | -0.052+/-0.749 | Large (+) Early<br>Large (-) Later |
| T Cell Specific Markers: |  |  |  |  |
| CD2 | all | NS | 0.039+/-0.000 | Negligible |
| CD3 | early | <0.002 | -0.984+/-0.000 | Large (-) Early |
|  | late | NS | 0.039+/-0.000 | Negligible Late |
| CD4 | *15 | <0.002 | -0.840+/-0.223 | Large (-) |
| CD8 | *I(2,5) | NS | 0.184+/-0.266 | Negligible |
| Both B and T Cell Markers: |  |  |  |  |
| CD45 | *20 | NS | -0.203+/-0.208 | Small |
| Clinical Features: |  |  |  |  |
| Splenomegaly | all | <0.002 | 0.984+/-0.000 | Large (+) |
| Anemia | all | <0.002 | 0.951+/-0.013 | Large (+) |
| Temperature | all | NS | 0.000+/-0.000 | No Effect |
| Hematologic Features: |  |  |  |  |
| Leukocytes/WBC | all | <0.002 | -0.984+/-0.000 | Large (-) |
| Lymphoblasts | all | <0.002 | 0.984+/-0.000 | Large (+) |
| Platelets |  | Variable | -0.249+/-0.443 | Variable (-) |
| Biochemical Markers: |  |  |  |  |
| LDH | all | <0.002 | 0.984+/-0.000 | Large (+) |
| MPO | all | <0.002 | 0.604+/-0.150 | Large (+) |
| Immunoglobulins: |  |  |  |  |
| IgA | all | <0.002 | 0.945+/-0.035 | Large (+) |
| IgG | all | <0.002 | -0.984+/-0.000 | Large (-) |
| IgM | all | <0.002 | -0.918+/-0.026 | Large (-) |

3. (TEL-AML1) fusion plus Family History (FHx) of Leukemia (Any): These data are summarized in Table 3

**Table 3:** Summary of the Cliff’s delta (d) effect size data for the combination of TEM-AML1 gene fusion plus Family Hixtory of Leukemia across all six Age cohorts.

| Age | TEM-AML1 Fusion | Splenomegaly | Anemia | Fever | CD10/Neprilysin | CD19 | CD2 | CD20 | CD22 | CD3 | CD4 | CD45 |
| --- | --- | --- | --- | --- | --- | --- | --- | --- | --- | --- | --- | --- |
| 0 | 0.984 | 0.984 | 0.984 | 0.000 | 0.984 | 0.984 | 0.039 | 0.984 | 0.984 | -0.984 | -0.984 | 0.039 |
| 2-3 | 0.984 | 0.984 | 0.945 | 0.000 | 0.984 | 0.984 | 0.039 | 0.984 | 0.984 | -0.984 | -0.984 | 0.000 |
| 5 | 0.984 | 0.984 | 0.945 | 0.000 | 0.984 | 0.984 | 0.039 | 0.984 | 0.984 | -0.984 | -0.984 | -0.354 |
| 10 | 0.984 | 0.984 | 0.945 | 0.000 | 0.984 | 0.984 | 0.039 | 0.984 | 0.984 | -0.039 | -0.984 | -0.394 |
| 15 | 0.984 | 0.984 | 0.945 | 0.000 | 0.984 | 0.984 | 0.039 | 0.984 | 0.984 | -0.039 | -0.984 | -0.394 |
| 20 | 0.984 | 0.984 | 0.945 | 0.000 | 0.984 | 0.984 | 0.039 | 0.984 | 0.984 | -0.039 | -0.984 | -0.551 |
| Average | 0.984 | 0.984 | 0.951 | 0.000 | 0.984 | 0.984 | 0.039 | 0.984 | 0.984 | -0.512 | -0.984 | -0.276 |
| 95% CI | 0.000 | 0.000 | 0.013 | 0.000 | 0.000 | 0.000 | 0.000 | 0.000 | 0.000 | 0.414 | 0.000 | 0.191 |
| Scale | -1 | -0.5 | 0 |  | 0.5 | 1 |  |  |  |  |  |  |
| Age | CD79a | CD8 | IgA | IgG | IgM | LDH | LeukocytesWBC | Lymphoblasts | MPO | Platelets | T Cell Receptor | TdT |
| 0 | 0.905 | -0.039 | 0.039 | -0.984 | -0.984 | 0.984 | -0.984 | 0.984 | 0.512 | -0.984 | 0.039 | 0.984 |
| 2-3 | 0.905 | 0.748 | 0.669 | -0.984 | -0.905 | 0.984 | -0.984 | 0.984 | 0.984 | -0.984 | 0.039 | 0.984 |
| 5 | 0.866 | 0.590 | 0.512 | -0.984 | -0.905 | 0.984 | -0.984 | 0.984 | 0.551 | 0.039 | 0.039 | 0.984 |
| 10 | -0.748 | -0.039 | 0.984 | -0.984 | -0.984 | 0.984 | -0.984 | 0.984 | 0.512 | -0.748 | -0.827 | 0.984 |
| 15 | -0.905 | 0.039 | 0.905 | -0.984 | -0.984 | 0.984 | -0.984 | 0.984 | 0.551 | -0.905 | 0.039 | 0.984 |
| 20 | -0.827 | -0.039 | 0.984 | -0.984 | -0.905 | 0.984 | -0.984 | 0.984 | 0.512 | 0.039 | 0.039 | 0.984 |
| Average | 0.033 | 0.210 | 0.682 | -0.984 | -0.945 | 0.984 | -0.984 | 0.984 | 0.604 | -0.590 | -0.105 | 0.984 |
| 95% CI | 0.754 | 0.288 | 0.294 | 0.000 | 0.034 | 0.000 | 0.000 | 0.000 | 0.150 | 0.396 | 0.283 | 0.000 |

3. (TEL-AML1) fusion plus Family History (FHx) of Leukemia (Any): These data are summarized in Table 3
| Genetic and Molecular Markers: | Age | p value | Cliff's d (±95%CI) | Effect size |
| --- | --- | --- | --- | --- |
| FG[TEL-AML1] | all | <0.002 | 0.984+/-0.000 | Large (+) |
| TdT | all | <0.002 | 0.984+/-0.000 | Large (+) |
| T Cell Receptor | all(10) | NS | -0.105+/-0.283 | Negligible |
| B Cell Specific Markers: |  |  |  |  |
| CD10/Neprilysin | all | <0.002 | 0.984+/-0.000 | Large (+) |
| CD19 | all | <0.002 | 0.984+/-0.000 | Large (+) |
| CD20 | all | <0.002 | 0.984+/-0.000 | Large (+) |
| CD22 | all | <0.002 | 0.984+/-0.000 | Large (+) |
| CD79a | all | <0.002 | 0.033+/-0.754 | Large (+)Early<br>Large (-) Late |
| T Cell Specific Markers: |  |  |  |  |
| CD2 | all | NS | 0.029+/-0.000 | Negligible |
| CD3 |  | Variable | -0.512+/-0.414 | Large (-) Early<br>No Effect Late |
| CD4 | all | <0.002 | -0.984+/-0.000 | Large (-) |
| CD8 | *2-5 | NS | 0.210+/-0.288 | Large (+)Early |
| Both B and T Cell Markers: |  |  |  |  |
| CD45 | most | NS | -0.276+/-0.191 | Small (-) |
| Clinical Features: |  |  |  |  |
| Splenomegaly | all | <0.002 | 0.984+/-0.000 | Large (+) |
| Anemia | all | <0.002 | 0.951+/-0.013 | Large (+) |
| Temperature | all | NS | 0.000+/-0.000 | No Effect |
| Hematologic Features: |  |  |  |  |
| Leukocytes/WBC | all | <0.002 | -0.984+/-0.000 | Large (-) |
| Lymphoblasts | all | <0.002 | 0.984+/-0.000 | Large (+) |
| Platelets | most | <0.002 | -0.590+/-0.396 | Medium (-) |
| Biochemical Markers: |  |  |  |  |
| LDH | all | <0.002 | 0.984+/-0.000 | Large (+) |
| MPO | all | <0.002 | 0.604+/-0.159 | Large (+) |
| Immunoglobulins: |  |  |  |  |
| IgA | *0 | <0.002 | 0.682+/-0.294 | Large (+) |
| IgG | all | <0.002 | -0.984+/-0.000 | Large (-) |
| IgM | all | <0.002 | -0.945+/-0.035 | Large (-) |

### Summary

Children with the TEL-AML1 fusion gene and a family history of leukemia exhibit significant hematologic and immunologic changes compared to the wild-type (WT) state. Notable findings include large increases in splenomegaly (Cliff’s delta = 0.984, p < 0.002) and lymphoblasts (Cliff’s delta = 0.984, p < 0.002), indicating large effect sizes. Additionally, there are significant increases in several B-cell markers such as CD10/Neprilysin, CD19, CD20, and CD22 (all with Cliff’s delta = 0.984, p < 0.002), highlighting heightened B-cell activity. Conversely, there are significant decreases in leukocyte counts (Cliff’s delta = −0.984, p < 0.002) and significant changes in immunoglobulins IgG and IgM, indicating large negative effects.

Other notable changes include a large decrease in early T-cell marker CD3 (Cliff’s delta = −0.512) and CD4 (Cliff’s delta = −0.984, p < 0.002). The overall pattern of increased immature lymphoid cells and decreased functional leukocytes suggests a predisposition to hematologic abnormalities. These findings underscore the significant impact of the TEL-AML1 fusion gene and FHx on hematologic and immunologic parameters in children, highlighting the need for close monitoring and early intervention in those with this genetic alteration.

The Link between a TEL-AML1 Gene Fusion and Early Childhood Hematologic Changes

The observed data align with known early indicators and risk factors for developing B-cell Acute Lymphoblastic Leukemia (B-ALL). The significant increase in lymphoblasts (Cliff’s delta = 0.984) and splenomegaly (Cliff’s delta = 0.984) are consistent with early leukemic changes. The heightened B-cell activity, as evidenced by increased CD19 levels, mirrors the overproduction of B-cells characteristic of B-cell ALL. Decreases in leukocyte counts, platelets, and certain immunoglobulins suggest early bone marrow stress and immune changes, which can precede the onset of leukemia. These data indicate a potential early disruption in hematologic and immunologic parameters, emphasizing the importance of vigilant monitoring and preventive care to mitigate the risk of progression to B-cell ALL in children with the TEL-AML1 fusion gene and a FHx of leukemia (16,17,18,19).

## Discussion

### 1. Summary of Key Findings

This virtual study utilized aiHumanoid simulations to explore the impact of the TEL-AML1 gene fusion with and without a family history of leukemia on childhood acute lymphoblastic leukemia (ALL). The results revealed significant hematologic and immunologic changes in children with these risk factors compared to unaffected/wild-type (WT) subjects. Children with a family history of leukemia exhibited marked increases in splenomegaly and lymphoblasts, along with heightened B-cell activity, evidenced by increased CD19 levels. Conversely, they showed significant decreases in leukocyte counts, platelets, and immunoglobulins (IgG and IgM), suggesting early bone marrow stress and immune changes consistent with a predisposition to hematologic abnormalities.

Based on the results data, the preleukemic state in children with either TEL-AML1 fusion, family history of leukemia, or both, appears to be predominantly B cell in nature. This conclusion is supported by the consistent increases in B cell markers (TdT, CD10, CD19, CD20, and CD22) and B cell activity, along with significant decreases in T cell markers during early stages which do not outweigh the B cell predominance.

### 2. Comparison with Existing Literature

Our findings align with previous studies identifying the TEL-AML1 fusion gene as a critical factor in ALL. The significant increase in B-cell activity markers and early leukemic changes observed are consistent with earlier research indicating that the TEL-AML1 fusion gene requires additional genetic alterations, or *Second Hits*, to cause acute leukemia [23,24,25]. The hematologic changes in children with a family history of leukemia corroborate studies that report a higher risk of developing ALL in these populations [20,21,27,35].

### 3. Implications for Clinical Practice

The identification of significant hematologic and immunologic markers provides potential for early diagnosis and monitoring of at risk children. Evidence of splenomegaly along with increased lymphoblasts, and B-cell markers (CD19) could serve as early indicators of leukemic changes, prompting closer surveillance and early intervention. The decreases in leukocyte (WBC) counts, platelets, and immunoglobulins highlight the need for vigilant monitoring to mitigate the risk of progression to overt leukemia [20,22,27]. Clinicians should consider integrating these findings into regular screening protocols for children with known risk factors.

### 4. Strengths and Limitations of the present study

This study’s primary strength lies in its innovative use of aiHumanoid simulations, enabling comprehensive longitudinal analysis without the limitations of attrition commonly seen in traditional studies [25,26]. However, the virtual nature of the study introduces limitations, such as potential biases in the simulated data and the need for validation in real-world cohorts [35,37,38,39]. Additionally, while the aiHumanoid simulations provide valuable insights, they may not fully capture the complexity of human biology [35].

### 5. Future Research Directions

Future research will investigate the impact of additional genetic alterations, or *Second Hits*, on the development and progression of ALL. This research will assess the effects of loss-of-function (LOF) mutations in Ikaros, p16, PAX5, and TEL LOH, as well as previous radiation exposure [29,31,32,33]. Further studies should validate these findings in clinical settings, involving larger and more diverse populations to enhance the generalizability of the results [30,32].

### 6. Conclusions

This study emphasizes the significant impact of the TEL-AML1 fusion gene and a familial history of leukemia on childhood ALL. The use of aiHumanoid simulations provides valuable insights into the genetic and immunologic changes associated with these risk factors, emphasizing the need for early diagnosis, vigilant monitoring, and tailored treatment strategies. The preleukemic state observed is predominantly B cell in nature, supported by significant increases in B cell markers (TdT, CD10, CD19, CD20, and CD22) and B cell activity. We intend to continue to explore the complex interactions between genetic alterations and the TEL-AML1 gene fusion to enhance our understanding and management of childhood ALL [21,35,36].

## Data Availability

The data is contained in the paper

## Appendix A: Reference list for Dx feature list used for ALL in children TEL-AML1 fusion gene

Zelent, A., Greaves, M. & Enver, T. Role of the *TEL-AML1* fusion gene in the molecular pathogenesis of childhood acute lymphoblastic leukaemia. *Oncogene* 23, 4275–4283 (2004). https://doi.org/10.1038/sj.onc.1207672

Splenomegaly:

Piscaglia, A. C., Rutigliano, L., Valente, M., & Rigante, D. (2020). Splenomegaly in Children and Adolescents. Frontiers in Pediatrics, 8, 581. https://doi.org/10.3389/fped.2020.00581

Anemia:

Bhatia, M., & Walters, M. C. (2014). Anemia in the pediatric patient. Blood, 124(18), 2783-2785. https://doi.org/10.1182/blood-2014-07-567741

Fever:

Chiappini, E., Bortone, B., Galli, L., & de Martino, M. (2017). Guidelines for the symptomatic management of fever in children: systematic review of the literature and quality appraisal with AGREE II. BMJ Open, 7(7), e015404. https://doi.org/10.1136/bmjopen-2016-015404

CD10/Neprilysin:

Shipp, M. A., Look, A. T. (1993). Hematopoietic differentiation antigens that are membrane-associated enzymes: cutting is the key!. Blood, 82(4), 1052-1070. https://doi.org/10.1182/blood.V82.4.1052.1052

CD19:

Park, J. H., Rivière, I., Gonen, M., Wang, X., Sénéchal, B., Curran, K. J., … & Sadelain, M. (2018). Long-term follow-up of CD19 CAR therapy in acute lymphoblastic leukemia. New England Journal of Medicine, 378(5), 449-459.

CD2:

Yang, J. J., Landier, W., Yang, W., Liu, C., Hageman, L., Cheng, C., … & Relling, M. V. (2015). Inherited NUDT15 variant is a genetic determinant of mercaptopurine intolerance in children with acute lymphoblastic leukemia. Journal of Clinical Oncology, 33(11), 1235.

CD20:

Thomas, D. A., O’Brien, S., Faderl, S., Garcia-Manero, G., Ferrajoli, A., Wierda, W., … & Kantarjian, H. M. (2006). Chemoimmunotherapy with a modified hyper-CVAD and rituximab regimen improves outcome in de novo Philadelphia chromosome–negative precursor B-lineage acute lymphoblastic leukemia. Journal of Clinical Oncology, 24(24), 3894-3902.

CD22:

Kantarjian, H., Thomas, D., Jorgensen, J., Jabbour, E., Kebriaei, P., Rytting, M., … & Garris, R. (2012). Inotuzumab ozogamicin, an anti-CD22–calecheamicin conjugate, for refractory and relapsed acute lymphocytic leukaemia: a phase 2 study. The Lancet Oncology, 13(4), 403-411.

CD3:

Pui, C. H., Evans, W. E. (2006). Treatment of acute lymphoblastic leukemia. New England Journal of Medicine, 354(2), 166-178.CD4

CD45:

Espinoza-Gutarra, M. R., Agarwal, P., Ferrer, L., Czader, M., & Dave, U. (2020). Relationship between CD45 Expression and Outcomes in B Lymphoblastic Leukemia/Lymphoma. Blood, 136 (Supplement 1), 24.

CD79a:

Pastore, S. I., Copetti, V., De Pieri, C., Radillo, O., Taddio, A., Ventura, A., & Tommasini, A. (2024). Childhood B-acute lymphoblastic leukemia: a genetic update. Experimental Hematology & Oncology, 3, 16.

CD8:

O’Dwyer, K. M. (2022). Optimal approach to T-cell ALL. Hematology, ASH Education Program.

IgA, IgG, and IgM in ALL in children

Kaczmarek, U., & Malicka, B. (2005). Oral mucositis and saliva IgA, IgG, and IgM concentration during anti-tumor treatment. Oral Oncology, 41(1), 98-105.

Hrones M, Tsang P. Acute lymphoblastic leukemia / lymphoma (2021). PathologyOutlines.com website. https://www.pathologyoutlines.com/topic/lymphnodesALL.html.

Parikh, S. A., Leis, J. F., Chaffee, K. G., Call, T. G., Hanson, C. A., Ding, W., … & Kay, N. E. (2014). Hypogammaglobulinemia in newly diagnosed chronic lymphocytic leukemia: Natural history, clinical correlates, and outcomes. Cancer, 121(17), 2883-2891.LDH:

Ross, A. D., & Abrahamson, E. (2011). A review of the use of serum lactate dehydrogenase measurement in patients presenting to the paediatric emergency department. Archives of Disease in Childhood, 96(Suppl 1), A872.

Leukocytes/WBC and Lymphoblasts:

Roberts, K. G. (2018). Genetics and prognosis of ALL in children vs adults. Hematology Am Soc Hematol Educ Program, 2018(1), 137–145.

MPO:

Varotto, E., Munaretto, E., Stefanachi, F., Della Torre, F., Buldini, B. (2022). Diagnostic challenges in acute monoblastic/monocytic leukemia in children. Frontiers in Pediatrics, 10

Platelets:

Bhojwani, D., & Pui, C. H. (2013). Relapsed childhood acute lymphoblastic leukaemia. The Lancet Oncology, 14(6), e205-e217

T Cell Receptor:

O’Dwyer, K. M. (2022). Optimal approach to T-cell ALL. Hematology Am Soc Hematol Educ Program, 2022(1), 197–205.

Terminal deoxynucleotidyl transferase (TdT):

O’Dwyer, K. M. (2022). Optimal approach to T-cell ALL. Hematology Am Soc Hematol Educ Program, 2022(1), 197–205

General Reviews of ALL/B-ALL in children

Al-Mahayri, Z. N., AlAhmad, M. M., & Ali, B. R. (2021). Long-Term Effects of Pediatric Acute Lymphoblastic Leukemia Chemotherapy: Can Recent Findings Inform Old Strategies? Frontiers in Oncology, 11

Roberts, K. G. (2018). Genetics and prognosis of ALL in children vs adults. Hematology, American Society of Hematology Education Program, 2018<u>(1), 137-145</u>

Targeting vulnerability in B-cell development leads to novel drug combination for leukemia (2024, April 8) retrieved 28 May 2024 from https://medicalxpress.com/news/2024-04-vulnerability-cell-drug-combination-leukemia.html

Talleur, Aimee C., Ching-Hon Pui, and Seth E. Karol. 2023. “What Is Next in Pediatric B-Cell Precursor Acute Lymphoblastic Leukemia” Lymphatics 1, no. 1: 34-44. https://doi.org/10.3390/lymphatics1010005

